# Phosphate levels and pulmonary damage in COVID-19 patients based on CO-RADS scheme: is there any link between parathyroid gland and COVID-19?

**DOI:** 10.1101/2020.08.25.20181453

**Authors:** Farshid Javdani, Shima Parsa, Heshmatollah Shakeri, Naser Hatami, Navid Kalani, Marzieh Haghbeen, Rahim Raufi, Alireza Abbasi, Pouyan Keshavarz, Seyyed Abbas Hashemi, Amin shafiee

## Abstract

**Background:** Preliminary studies of COVID-19 have provided some evidence that electrolyte disturbances may also be present in patients. In this study we aimed to evaluate the role of the arrival electrolytes and symptoms in prediction of Lung damage in CT scan based on the CO-RADS system.

**Methods:** This was a retrospective cross-sectional analytical study. We included patients with laboratory confirmed COVID-19 infection, June 15 to July 7. Patients were included in study if there were no previous history of kidney disease. Demographic, clinical characteristics, laboratory findings, and CO-RADS High-resolution computed tomography (HRCT) of lung report were collected. Univariate logistic regression was employed first to identify the effective, correlated items. All statistics were performed with SPSS version 18.0.

**Results:** In the current study, 36 (20 male- 16 female) patients with mean age of the 54.7±17.5 years old were studied. Most common symptom at the arrival was the Fever (52.8%), followed by Fatigue (18%), and dyspnea (44.4%). Computed tomography assessment revealed CO-RADS 2 in 4 (11.1%) patients, CORADS 3 in 1 (2.8%), CO-RADS 4 in 20 (55.6%), and CO-RADS 5 in 11 (30.6%) patients. In the comparison with the study groups based on the HRCT status (CO-RADS II,III vs. CO-RADS IV,V), patients with severe HRCT damage had significantly lower level of Phosphorus (*P* < 0.01). Univariate logistic regression analysis showed that only one factors was associated with HRCT damage status (Phosphorus, *P*=0.040). Phosphorus upper than 4.5 was associated with better HRCT results with OR ratio of 3.71 (X2(1)=5.69; p=0.017).

**Conclusion:** Our study illustrates that higher phosphate levels may be associated with better CT scan of lung outcomes in COVID-19; while hypophosphatemia is associated with severe lung injuries. This could help clinicians to manage hospitalized patients and may link the COVID-19 and parathyroid gland.

## Introduction

A new infectious disease, caused by coronavirus 2 (SARS-CoV-2), was discovered in Wuhan, China in December 2019. The rapid emergence of COVID-19 in Wuhan, China, to all around the world has resulted in thousands of deaths worldwide. However, many infected patients show mild symptoms such as the common cold and recover quickly, there are extremely increasing number of deaths due to COVID-19. Preliminary case reports and cohort studies have described many clinical features of patients with coronavirus 2019 (COVID-19). Preliminary studies of COVID-19 have provided some evidence that electrolyte disturbances may also be present in patients, including sodium, potassium, chloride, and calcium abnormalities (1,2). No specific treatment is currently available, and current management includes supportive medical care (3). Electrolyte disturbances can have important implications not only for patient management (4), but also for the identification of potential pathophysiological mechanisms underlying COVID-19, which in turn can lead to new treatment opportunities. However, limited and heterogeneous sample sizes can be seen in the report of electrolyte interpretation in studies to date. Therefore, in order to compare the analysis in this study, we evaluate the electrolyte properties in the initial trials of patients with confirmed COVID-19 (PCR) infection in Jahrom city of Iran and the relationship with the involvement of patients in HRCT.

## Method

### Study design

This was a retrospective cross-sectional analytical study. We admitted patients with COVID-19 infection who were transferred from clinics to designated hospitals for the treatment of specific infectious diseases (June 15 to July 7). The study design of this study was approved by the ethics committee of Jahrom University of Medical Sciences (code of IR.JUMS.REC.1398.130) and the written informed consent was obtained from the patients due to the retrospective nature of the study. Inclusion criteria of study was Patients who were referred to the hospital with positive PCR test for COVID-19, if there were no previous history of kidney disease (due to possible effect on electrolytes). Exclusion criteria was Patient dissatisfaction to participate in the stud.

### Data collection

At the patient arrival at emergency department of COVID-19 special ward, 5 cc blood was taken for electrocytes analysis and blood sugar was assessed by glucometer. History and physical examination taking was performed by physician. Demographic variables were also recorded. CO-RADS having 5 levels was the classification method used to address severity of lung injury in COVID-19 patients. Category I of CO-RADS indicates a relatively small level of suspicion to COVID-19 pulmonary involvement; category V of CO-RADS implies a very strong level of suspicion of COVID-19 pulmonary involvement based on standard CT findings like ground-glass opacities (5). CT scan was performed at the arrival of patients.

### Statistical analysis

Continuous variables are presented as the mean ± standard derivation (SD), and a Student’s *t*-test was used to determine whether there was a significant difference between the two groups based on the HRCT results (Table 1). Continuous variables with significant differences were transformed into dummy variables. To examine the effect of demographic factors on the HRCT lung damage, univariate logistic regression was employed first to identify the effects, correlated items. All statistics were performed with SPSS version 18.0, and the level of significance was set at *P* < 0.05.

**Table 1.** electrolytes comparison between CO-RADS II, III vs. IV, V

|  |  | mean | STD | P** | Std. Error Difference | 95% Confidence Interval of the Difference |  |
| --- | --- | --- | --- | --- | --- | --- | --- |
|  |  |  |  |  |  | Lower | Upper |
| Mg | CO-RADS II, III | 1.8 | 0.07 | 0.35 | 0.1 | -0.29 | 0.11 |
|  | CO-RADS IV, V | 1.89 | 0.21 |  | 0.05 | -0.2 | 0.01 |
| Phosphate | CO-RADS II, III | 5.36 | 1.63 | 0.002 | 0.59 | 0.74 | 3.12 |
|  | CO-RADS IV, V | 3.43 | 1.15 |  | 0.76 | -0.06 | 3.93 |
| BUN | CO-RADS II, III | 16.6 | 11.76 | 0.72 | 5.05 | -12.08 | 8.44 |
|  | CO-RADS IV, V | 18.42 | 10.29 |  | 5.57 | -16.12 | 12.48 |
| Creatinine* | CO-RADS II, III | 2.1 | 2.03 | 0.05 | 0.4 | 0 | 1.64 |
|  | CO-RADS IV, V | 1.28 | 0.5 |  | 0.91 | -1.69 | 3.33 |
| Potassium | CO-RADS II, III | 3.92 | 0.36 | 0.45 | 0.27 | -0.77 | 0.35 |
|  | CO-RADS IV, V | 4.13 | 0.59 |  | 0.19 | -0.65 | 0.23 |
| Na | CO-RADS II, III | 136.2 | 1.64 | 0.7 | 22.68 | -54.79 | 37.39 |
|  | CO-RADS IV, V | 144.9 | 50.1 |  | 9.03 | -27.13 | 9.72 |
| Albumin | CO-RADS II, III | 3.16 | 0.5 | 0.23 | 0.19 | -0.61 | 0.15 |
|  | CO-RADS IV, V | 3.39 | 0.37 |  | 0.23 | -0.84 | 0.39 |
| Blood sugar | CO-RADS II, III | 129.8 | 46.62 | 0.46 | 18.55 | -23.92 | 51.46 |
|  | CO-RADS IV, V | 116.03 | 37.26 |  | 21.9 | -43.01 | 70.54 |
|  | V |  |  |  |  |  |  |

\*non-parametric, \*\*two-tailed P-value

## Results

36 patients were evaluated in this study. There were 20 male and 16 female patients with a mean age of 54.7±17.5 years old. The most common symptom at the arrival was the Fever (52.8%), followed by Fatigue (18%), and dyspnea (44.4%). Computed tomography Assessment Scheme revealed CO-RADS 2 in 4 (11.1%) patients, CO-RADS 3 in 1 (2.8%),CO-RADS 4 in 20 (55.6%), and CO-RADS 5 in 11 (30.6%) patients. In comparison with the study groups based on the HRCT status (mild, severe), patients with severe HRCT damage had a significantly lower level of Phosphate (*P* < 0.01, Table 1). Phosphate was then transformed into a dummy variable (Table 2) and was used in the univariate logistic regression analysis.

**Table 2.**
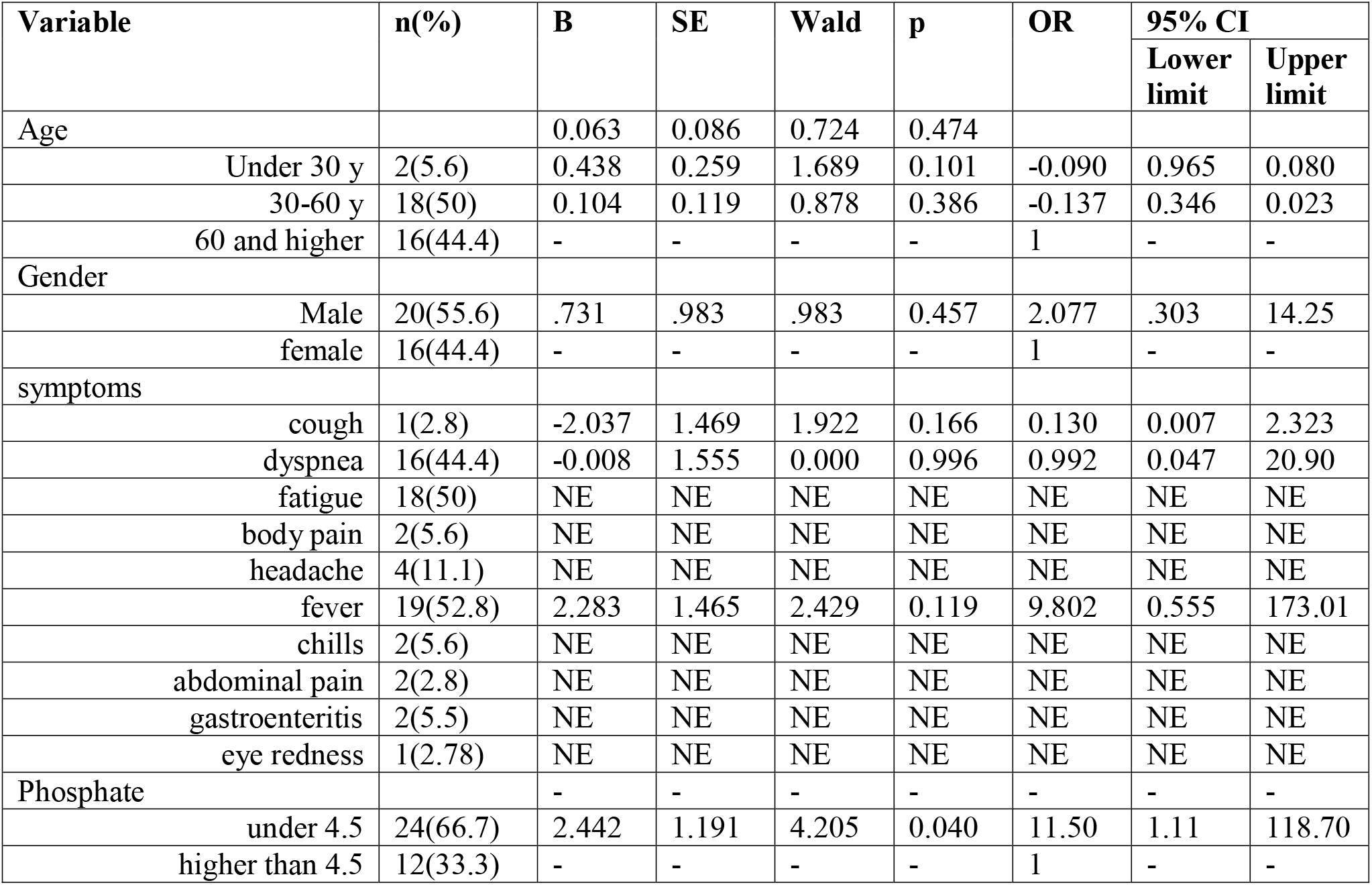
univariate logistic regression analysis for HRCT damage status

To carry out a univariate logistic regression analysis, HRCT damage status was set as the dependent variable, and the independent variables consisted of 13 factors including Age, Gender, cough, dyspnea, fatigue, body pain, headache, fever, chills, abdominal pain, gastroenteritis, eye redness, Phosphorus. Univariate logistic regression analysis showed that only one factor was associated with HRCT damage status (Phosphate, *P*=0.040). So, no further Multivariate analysis was conducted (table 2).

Phosphor upper than 4.5 was associated with better HRCT results with OR of 3.71 (X2(1)=5.69; p=0.017). As shown in figure 1, there were significant differences in serum level of phosphate in patients with CO-RADS IV and V versus patients with CO-RADS II (P<0.05).

**Figure 1.**
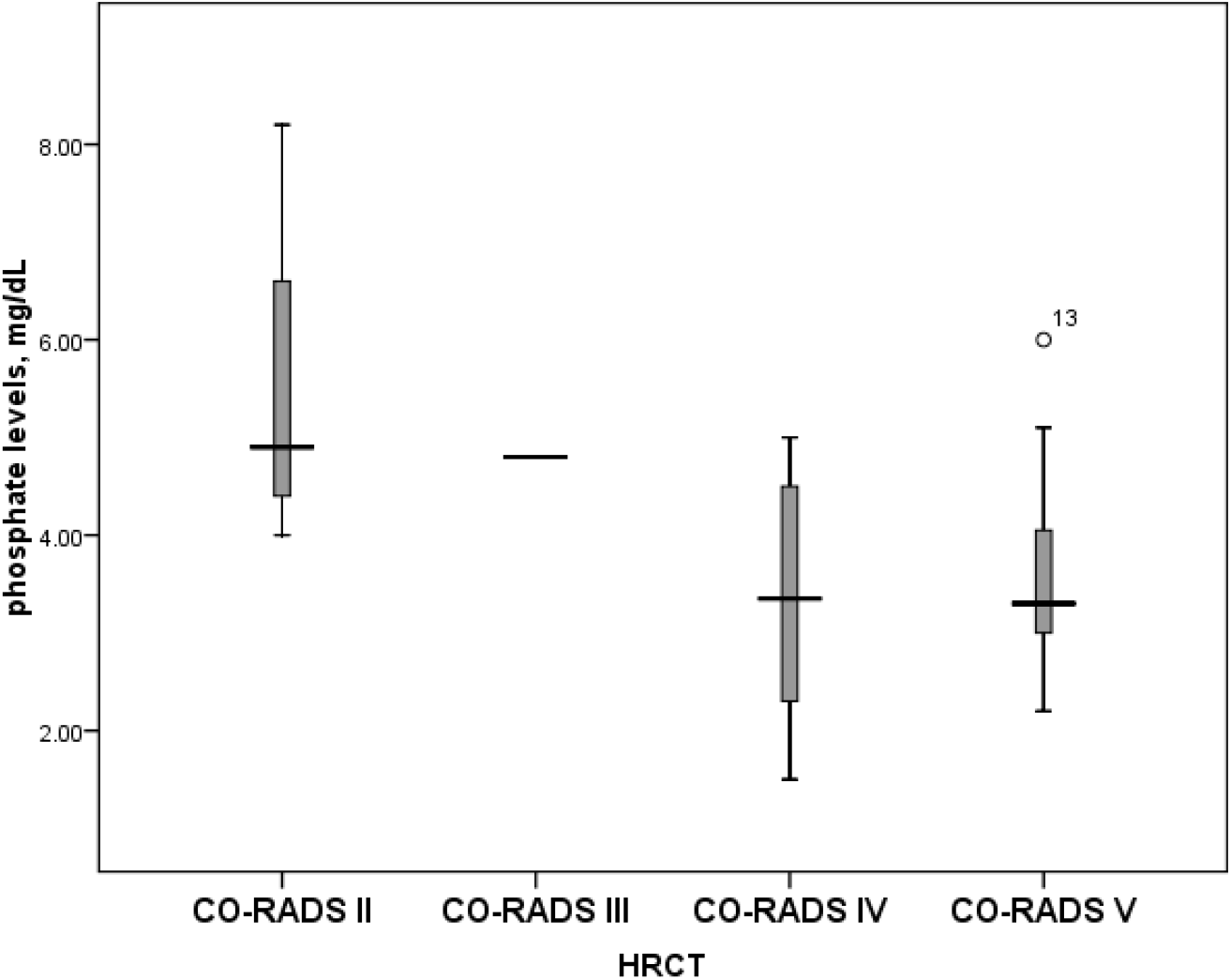
phosphate levels in each group of CO-RADS schemes

## Discussion

Our study revealed that higher phosphate levels may be associated with better CT scan of lung outcomes in COVID-19; while hypophosphatemia is associated with severe lung injuries. To our knowledge, it’s the first report of COVID-19 patients showing the possible protective effect of phosphate level of serum on lung injury caused by SARS-COV2 virus. There are some previous studies showing possible role of the hypophosphatemia in severity of disease. Kormann et al. (6) study evaluating 42 laboratory-confirmed COVID-19 patients, compared phosphate serum levels of patients admitted in ICU versus other patients admitted in medical department wards. Their result didn’t reveal any significant difference between the Baseline serum phosphate of these groups of patients. There were Hyperphosphataemia was just seen in 2 (5%) patients who were admitted in ICU while no patient in the medical department had hypophosphatemia. But in our study, there were 4 patients with Hyperphosphataemia among severely damaged lungs and 5 subjects among mild HRCT lung damage group. On the other hand, 7 patients had hypophosphatemia in our study and all were experiencing severe lung injury. Arenas et al. (7) study showed hypophosphatemia in renal patients with confirmed COVID-19 in comparison of renal patients who were negative for the COVID-19 PCR test. Our study is in contrast with Arenas et al. as we observed a wide range of serum phosphate levels in our patients with a reasonable association with HRCT results. They suggested hypophosphatemia as a potential risk factor of COVID-19; while it could be a reflection of malnutrition. But in our study phosphate levels were assessed in the association of COVID-19 severity and comparison of our results would not be possible as also their study sample were renal patients. As we know renal patients may show severe alternations in phosphate levels due to renal diseases and hemodialysis processes. On other hand hyperphosphatemia due to kidney inability of phosphate excretion is frequent in renal patients (8) and the hypothesis of Arenas et al. for hypophosphatemia association with COVID-19 make sense, as we expect up to 90% of renal patients to experience hyperphosphatemia (9), but further shreds of evidence are needed. Another study showed normal ranges of phosphate in 14,712 COVID-19 patients, with no differences among male and female cases (10). While their data was collected from multiple centers medical recorders through the TriNetX network and no information was addressed about the time of the laboratory analysis, their results may be biased by the different time point of laboratory results recording based on the patient course of the disease. But we assessed laboratory results at the arrival of patient to the Emergency department.

Phosphorus is one of the most important ions inside and outside the cell due to its role in physiological processes, especially muscle contraction and adenosine triphosphate production (11). Therefore, hypophosphatemia is one of the most important electrolyte disorders in special wards (12). The main causes of hypophosphatemia are diffuse infection, trauma, receiving volumetric agents, malnutrition syndrome, acid and base disorder, and some drugs, as well as conditions such as major surgery, hypoglycemia, and osmotic diuresis, parathyroidectomy and thyroidectomy, and malnutrition. (13). This complication is very important in patients under mechanical ventilation due to the effect of hypophosphatemia on muscle contraction (14); Weak muscle contraction leads to increased duration of mechanical ventilation and difficulty separating (15).

On the other hand, the findings of this study may be in line with researches looking for vitamin D benefits for COVID-19 patients. Calcitriol is the biologically active form of vitamin D, which is the main hormone that regulates calcium and phosphorus metabolism, and its blood concentration is strongly regulated by parathyroid hormone, calcium, and phosphorus.

## Conclusion

This study facing various limitations like doesn’t have a follow up for the patient treatment outcome and a low number of samples is showing that higher serum levels of phosphate are associated with milder involvement of lung in COVID-19 patients. The strength of this study is suggesting possible benefits of hyperphosphatemia for COVID-19 patients. But these results may have been biased by other confounding factors that weren’t assessed in our study as well as malnutrition. So further studies are needed in this area of research.

## Data Availability

all data referred to in the manuscript are available in manuscript.

## Acknowledgment

We would like to thank the Clinical Research Development Unit of Peymanieh Educational and Research and Therapeutic Center of Jahrom University of Medical Sciences for providing facilities for this work.

## Authors Contributions

All the authors met the criteria of authorship based on the recommendations of the international Committee of Medical Journal Editors.

## References

1. Sun J, He WT, Wang L, Lai A, Ji X, Zhai X, Li G, Suchard MA, Tian J, Zhou J, Veit M. COVID-19: epidemiology, evolution, and cross-disciplinary perspectives. Trends in Molecular Medicine. 2020 Mar 21.

2. Rothan HA, Byrareddy SN. The epidemiology and pathogenesis of coronavirus disease (COVID-19) outbreak. Journal of autoimmunity. 2020 Feb 26:102433.

3. Jin Y, Yang H, Ji W, Wu W, Chen S, Zhang W, Duan G. Virology, epidemiology, pathogenesis, and control of COVID-19. Viruses. 2020 Apr;12(4):372.

4. Lippi G, South AM, Henry BM. Electrolyte imbalances in patients with severe coronavirus disease 2019 (COVID-19). Annals of Clinical Biochemistry. 2020 May;57(3):262–5.

5. Prokop M, van Everdingen W, van Rees Vellinga T, van Ufford JQ, Stöger L, Beenen L, Geurts B, Gietema H, Krdzalic J, Schaefer-Prokop C, van Ginneken B. CO-RADS-A categorical CT assessment scheme for patients with suspected COVID-19: definition and evaluation. Radiology. 2020 Apr 27.

6. Kormann R, Jacquot A, Alla A, Corbel A, Koszutski M, Voirin P, Garcia Parrilla M, Bevilacqua S, Schvoerer E, Gueant JL, Namour F. Coronavirus disease 2019: acute Fanconi syndrome precedes acute kidney injury. Clinical Kidney Journal. 2020 Jun 8.

7. Arenas MD, Crespo M, Pérez-Sáez MJ, Collado S, Redondo-Pachón D, Llinàs-Mallol L, Montero MM, Villar-García J, Arias-Cabrales C, Barbosa F, Buxeda A. Clinical Profiles in Renal Patients with COVID-19. Journal of Clinical Medicine. 2020 Aug;9(8):2665.

8. Shaman AM, Kowalski SR. Hyperphosphatemia management in patients with chronic kidney disease. Saudi Pharmaceutical Journal. 2016 Jul 1;24(4):494–505.

9. Elseviers M, De Vos JY. The use of phosphate binders: data from contributors to the European Practice Database. Journal of renal care. 2009 Mar;35:14-8.

10. Alkhouli M, Nanjundappa A, Bates MC, Bhatt DL. Sex Differences in Case Fatality Rate of COVID-19: Insights From a Multinational Registry. InMayo Clinic Proceedings 2020 Aug 1 (Vol. 95, No. 8, pp. 1613-1620). Mayo Foundation for Medical Education and Research.

11. Uribarri J. Phosphorus homeostasis in normal health and in chronic kidney disease patients with special emphasis on dietary phosphorus intake. InSeminars in dialysis 2007 (Vol. 20, No. 4, pp. 295-301).

12. Suzuki S, Egi M, Schneider AG, Bellomo R, Hart GK, Hegarty C. Hypophosphatemia in critically ill patients. Journal of critical care. 2013 Aug 1;28(4):536-e9.

13. Gaasbeek A, Meinders AE. Hypophosphatemia: an update on its etiology and treatment. The American journal of medicine. 2005 Oct 1;118(10):1094–101.

14. Alsumrain MH, Jawad SA, Imran NB, Riar S, DeBari VA, Adelman M. Association of hypophosphatemia with failure-to-wean from mechanical ventilation. Annals of Clinical & Laboratory Science. 2010 Mar 20;40(2):144–8.

15. Zhao Y, Li Z, Shi Y, Cao G, Meng F, Zhu W, Yang GE. Effect of hypophosphatemia on the withdrawal of mechanical ventilation in patients with acute exacerbations of chronic obstructive pulmonary disease. Biomedical reports. 2016 Apr 1;4(4):413–6.

